# A Supervised Text-Embedded Transformer Matching Model to Detect Fall Injuries in Medicare Data

**DOI:** 10.64898/2026.07.29.26359258

**Authors:** Michael Kane, Erich J. Greene, Denise Esserman, Nancy K. Latham, Lillian C. Min, David A. Ganz

## Abstract

**Objective:** To develop and validate a supervised text-embedded transformer matching model to identify fall injuries in Medicare data, and evaluate the model’s performance -- alongside a validated rule-based algorithm-- against “ground truth” from an external reference standard (self-reported fall injuries leading to medical attention).

**Materials and Methods:** Text embeddings of ICD-10-CM and CPT codes in Medicare claims/encounters from participants in the Strategies to Reduce Injuries and Develop Confidence in Elders (STRIDE) trial served as model inputs. Trained on annotated claims/encounters occurring within +/- one month of self-reported fall injuries leading to medical attention, the transformer model generated a continuous 0-1 probability that each claim/encounter was for a fall injury. The model was then applied to all claims/encounters in STRIDE and compared alongside the rule-based algorithm to the external reference standard.

**Results:** The model achieved an area under the curve (AUC) of > 0.96 against annotated claims/encounters in 9 out of 10 holdout folds and 0.85 in the remaining fold. In the full STRIDE dataset, the model achieved a peak AUC of 0.86 (95% CI, 0.84-0.87) against the external reference standard, with results comparable to the rule-based algorithm.

**Discussion:** Relative to rule-based approaches, which typically generate binary outcomes, the continuous event probability generated by the transformer model could support clinical endpoint adjudication, with high-probability predictions treated as events, moderate-probability predictions being adjudicated, and low-probability predictions treated as non-events.

**Conclusion:** A text-embedded transformer model identified fall injuries with comparable accuracy to a rule-based algorithm, demonstrating “proof of concept” for use in endpoint adjudication.

## BACKGROUND AND SIGNIFICANCE

Clinical trials and epidemiologic studies can potentially save time and resources by using routinely collected administrative data (e.g., from healthcare claims or encounters) to ascertain acute clinical outcomes, such as falls, myocardial infarction, or stroke. Using routinely collected data avoids the need for separate data collection workflows (e.g., calling patients by phone to ascertain outcomes or requiring human review of the full text of medical records). Avoiding separate workflows can reduce burden on patients (thus increasing participation in research and reducing loss to follow-up). The advantages of using administrative data are magnified particularly in large, pragmatic multi-center studies in which thousands of individuals may be enrolled across different healthcare systems, or to measure the effects of population-wide programs within large healthcare systems. In these large studies, administrative data that accrue at the level of an insurer that includes all participants and cover all locations where healthcare may be received (e.g., Medicare in the case of individuals age 65 and older in the US) may simplify data collection and analysis relative to other routinely collected data sources such as electronic health records.^1^

However, routinely collected data have certain limitations, including that they were not collected with the level of accuracy and detail typical of data collected for research purposes.^2^ Administrative data, in particular, may suffer from inaccurate coding, unless an incentive exists to code properly. Diagnostic accuracy may be lower than in data collected expressly for the purpose of research. As a result, algorithms that use routinely collected data may vary in how well they capture the outcomes of interest, with variable sensitivity and specificity,^3^ potentially resulting in misclassification of the outcome and a reduction in statistical power. Thus, whether administrative data can substitute for data collected via traditional research methods is an empirical question.

Historically, algorithms to identify outcomes in administrative data have been rule-based, i.e., they follow rules that require the presence (or absence) of specific diagnostic and procedural codes in administrative datasets to identify an outcome. This approach typically produces a binary result: the outcome is either present or absent. One limitation of this approach is that it is inflexible to variations in coding practices and typically cannot identify a range of probabilities of events occurring (e.g., possible or probable events). Such information could be useful for “hybrid” outcome ascertainment approaches that supplement routinely collected data with additional adjudication of events that cannot be definitively identified from the routinely collected data alone. We hypothesized that a supervised transformer matching model that used large language model embeddings of the text descriptions of diagnostic and procedural codes as inputs, could offer better performance than an equivalent rule-based algorithm,^4,5^ while offering a more flexible platform for identifying acute clinical outcomes in administrative data.

Here, we describe the development and validation of the transformer model with a use case focused on identifying fall injuries in Medicare data. Our work proceeded in two phases: 1) model development and validation on a subset of available data that was annotated; and 2) model deployment in the full dataset, with comparison alongside the existing rule-based algorithm against an external reference standard (self-reported fall injuries leading to medical attention).

## MATERIALS AND METHODS

### Overview

This analysis uses data from the Strategies to Reduce Injuries and Develop Confidence in Elders (STRIDE) study, a clinical trial of a multifactorial fall injury prevention intervention compared with enhanced usual care.^6^ Details on linkage of the STRIDE study to Medicare data, validation of the rule-based algorithm, and the external reference standard are described in detail in a previous publication.^4^ This study was approved by Massachusetts General Brigham IRB, as an amendment to protocol 2015P000693.

### Data and Embeddings

In this analysis, each sample comprises a variable-length set of ICD-10-CM codes and Current Procedural Terminology (CPT) codes contained within a claim billed to traditional Medicare or an encounter from Medicare Advantage for a given patient. (In traditional Medicare, each claim submitted by a provider or facility represents a bill that is then paid by Medicare. In contrast, in Medicare Advantage, Medicare pays health plans a lump sum to manage all the care of each beneficiary; encounters are records of care received by patients but are not generated for billing purposes.) Notably, the number of ICD-10-CM and CPT codes per claim or encounter can vary quite widely across samples, reflecting real-world heterogeneity in clinical documentation. These codes serve as independent variables, capturing diagnostic and procedural information.

### Phase 1: Transformer Model Development and Validation

For model development and validation, the dependent variable is a binary indicator denoting whether each claim or encounter in question matched a study participant’s self-reported fall injury leading to medical attention. The determination of a match was made in the following way: first, we pulled all claims or encounters whose dates fell within one month in either direction of the date of the patient’s self-reported fall injury leading to medical attention. These claims or encounters constituted the sample that was annotated and the universe of data used for fitting the transformer model. Self-reported fall injuries leading to medical attention (n=2156), which were used to support the annotation process, derived from structured patient-reported outcome measures collected as part of the STRIDE study. These structured reports contained information about the date of the injury, the level of care received (i.e., overnight hospital stay, emergency department visit, doctor visit, and/or visit to another facility), and the type of injury (i.e., fracture, dislocation, head injury, cut with bleeding, strain or sprain, bruising or swelling, and/or other injury). For fractures, the body site was also reported. To create the binary indicator for presence or absence of a match for each claim or encounter, a geriatrician (DAG) reviewed each self-reported event together with the claims or encounters that were in chronological proximity. Within the claim or encounter, the reviewer was shown up to 25 ICD-10-CM diagnosis codes, up to 12 ICD-10-CM external cause of injury codes, and up to 12 health care common procedure coding system (HCPCS) codes, including CPT codes. If a claim appeared to match to the self-report data (with respect to containing ICD-10-CM and/or CPT codes that were, in the judgment of the reviewer, highly likely to reflect a claim or encounter for the injury in question, based on date, injury type, level of care received, and body site, if applicable), that claim was recorded as a match. Claims were recorded as a match regardless of whether they reflected initial care for the injury in question (e.g., in the emergency department) or subsequent care for that injury (e.g., in a skilled nursing facility).

For each sample, text descriptions of both the ICD-10-CM and CPT codes were embedded using a compressed large language model (LLM) approach, as detailed in our prior work.^7^ Briefly, this method leverages a fine-tuned LLM to produce high-dimensional vector representations of code descriptions, followed by dimensionality reduction via an autoencoder to yield compact embeddings (e.g., 1000 dimensions in the dataset used here). This process preserves semantic relationships among codes, such as hierarchical structures (e.g., chapters and blocks) and contextual similarities, which are critical for downstream tasks like fall injury prediction. For instance, codes related to musculoskeletal injuries or gait disturbances are embedded in proximity, facilitating the capture of fall-related patterns. These embeddings mitigate the challenges of high-dimensional, sparse categorical data in healthcare claims and encounters, enabling models to infer latent relationships.

The annotated dataset was preprocessed by concatenating each ICD-10-CM and CPT embedding (which was of similar length). For a single sample, which consisted of the ICD-10-CM and CPT codes in a given claim or encounter, two binary values were fused to each embedding, one indicating that the sample was an ICD-10-CM code (as opposed to a CPT code) and another indicating whether it was the first ICD-10-CM code in the sequence (the first code often indicates the primary reason for the service provided). The resulting vector was then stacked and padded to a maximum number of ICD-10 and CPT codes in the samples to create matrices of uniform size (41 by 1002).

### Phase 1 Model Architecture (Figure 1)

To estimate the dependent variable, we developed a fused transformer model implemented in R^8^ using the torch^9^ and luz^10^ packages. The architecture is designed to handle variable-length sequences of multimodal embeddings (ICD-10-CM and CPT), fusing them into a unified representation for binary classification.

**Figure 1.**
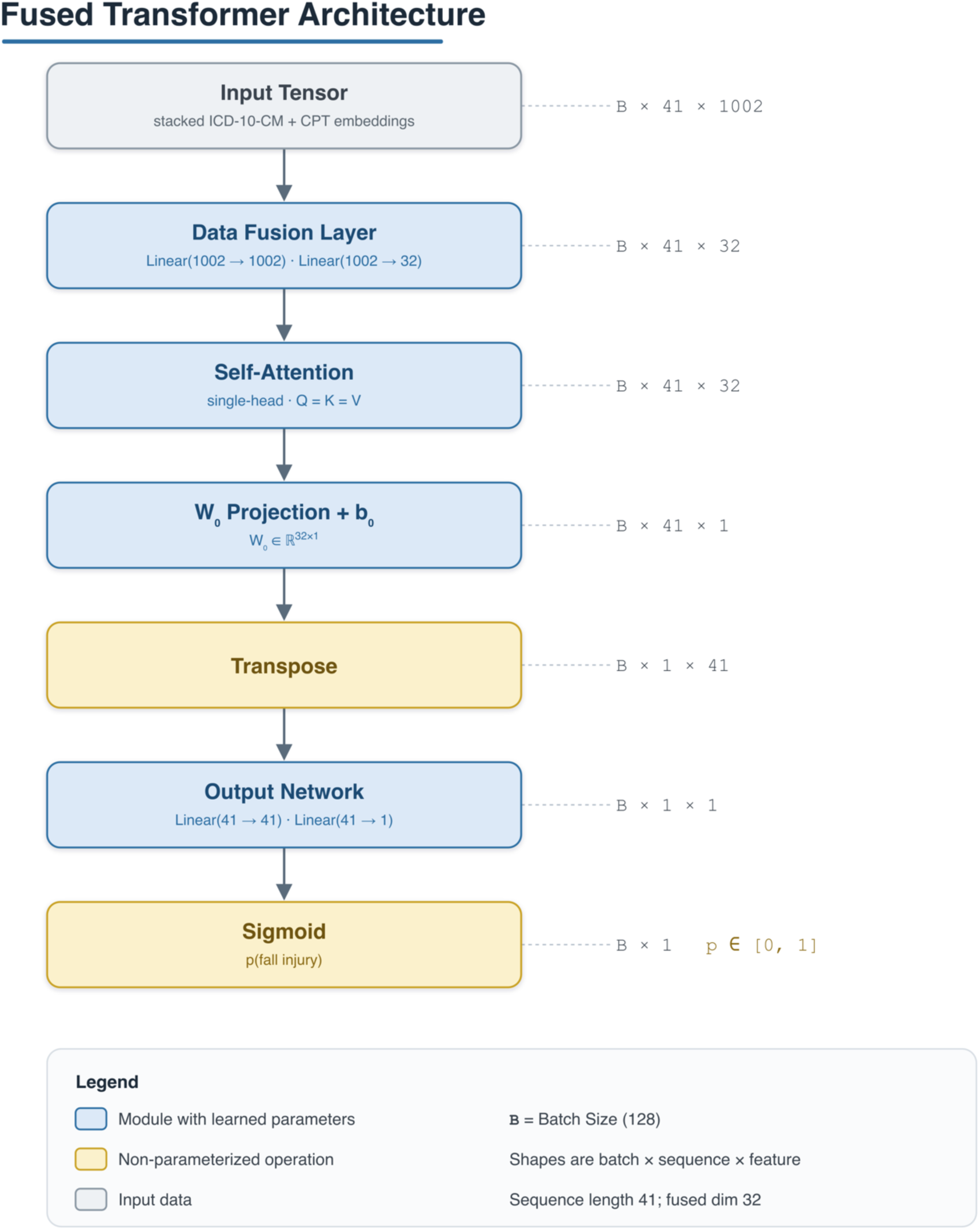
Transformer model architecture. Figure 1 shows the fused transformer model’s architecture. Input embeddings (1000-dimensional) are projected to a 32-dimensional fused space, processed by single-head self-attention, and reduced to a scalar prediction via a learned output projection. Tensor shapes are annotated at each stage. Blue boxes connote modules where parameters are learned; yellow boxes are non-parameterized operations.

The model initializes with the following hyperparameters: input sequence length (e.g., 41, based on empirical maximums), input embedding dimension (1002), and fused embedding dimension (32). The core components are:

- Data Fusion Layer: A sequential neural network comprising two linear layers. The first projects the input embeddings to an intermediate space (maintaining the original dimension), and the second reduces it to the fused dimension. This step integrates ICD-10-CM and CPT features, allowing the model to learn cross-modality interactions early in the pipeline.
- Self-Attention: A single-head attention mechanism^11^ processes the fused embeddings. Using batch-first configuration, it computes self-attention across the sequence, where queries, keys, and values are all derived from the fused inputs. This enables the model to weigh relevant codes (e.g., those indicative of balance issues or prior injuries) while attending to the entire context within the claim or encounter.
- Output Projection: The attention output is matrix-multiplied with a learnable weight matrix *W*_0_(initialized uniformly in [-0.001, 0.001]) and added to a bias *b*_0_(initialized in [-0.001, 0.001]). The result is transposed and projected through a two-layer learned linear network that reduces the sequence dimension to a scalar, then passed through a sigmoid activation to produce a probability of fall injury reporting (range: [0, 1]).

More formally, for input tensor *x* ∈ ℝ*^B^*^×*S*×*D*^ (where *B* is batch size, *S* is sequence length, *D* is input dimension), the forward pass is:

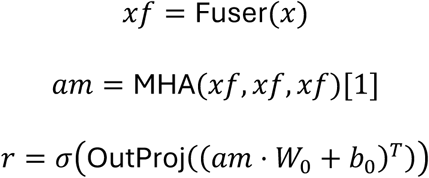

where *xf* refers to the fused input data, *am* is the attention matrix, MHA()[1] refers to the first output of the multi-head attention function, *r* is the estimate of a match on the inverse logit scale, *σ* is the sigmoid function, the output projection (OutProj) is a two-layer learned linear projection from the sequence dimension to a scalar, and superscripted *T* refers to the matrix transpose.

### Phase 1 Training and Evaluation (Figure 2)

The model was trained using a custom binary cross-entropy loss:

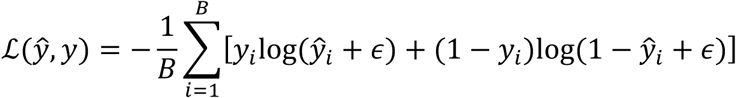

**Figure 2.**
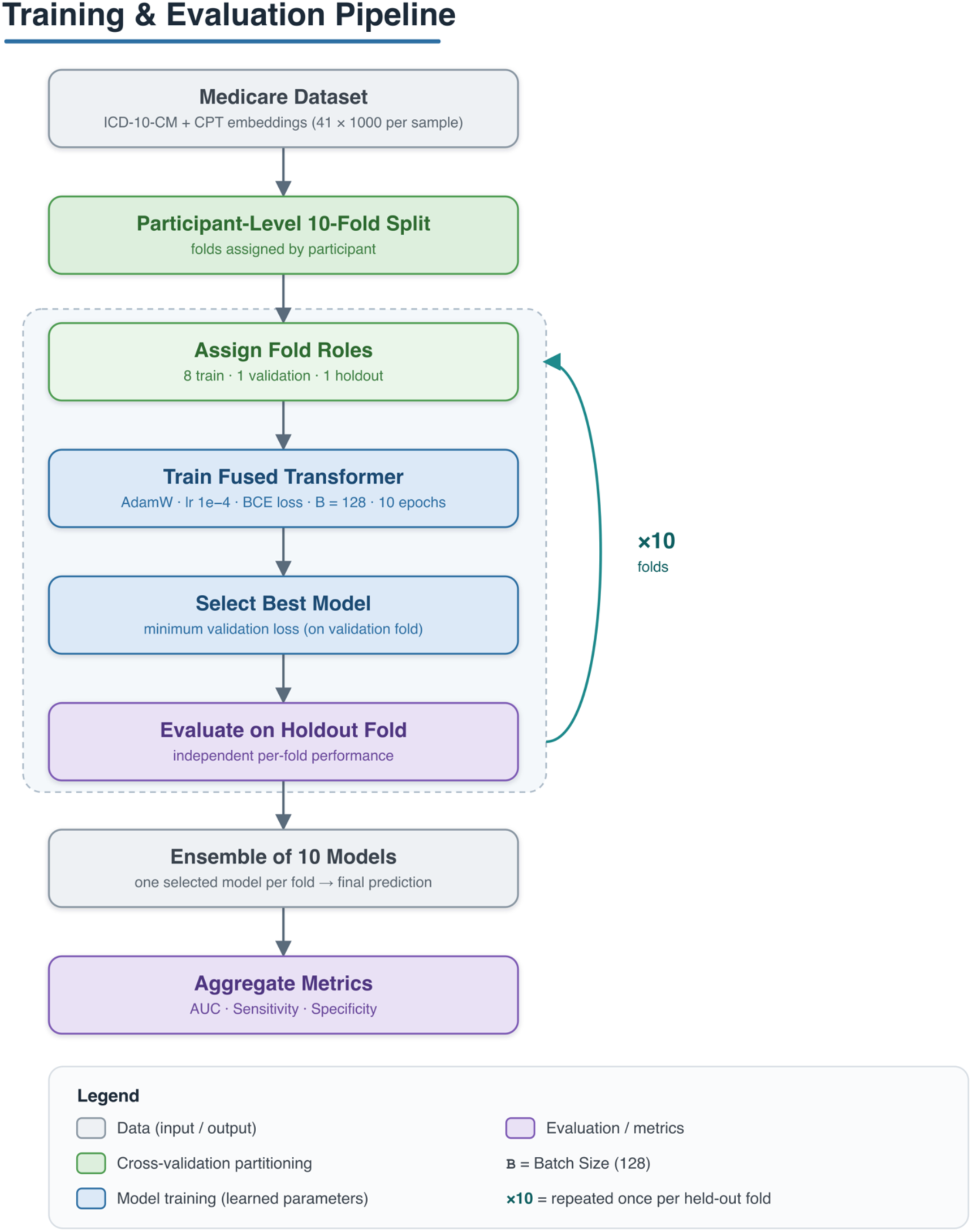
Training and Evaluation Pipeline Figure 2 shows the 10-fold cross-validation training pipeline. For each iteration, 8 folds are used for training, 1 for validation (model selection), and 1 as a holdout for independent evaluation. The 10 best models form an ensemble for final prediction.

with *ε* = 10^−6^ for numerical stability. Optimization employed AdamW^12^ with a learning rate of 10^−4^. Data were partitioned into 10 folds at the participant level to create an ensemble of 10 models. For each fitting, one fold was used as a holdout (performance evaluation), one fold was used for validation, and the remaining eight were used for training. Training ran for 10 epochs per fold, using batch size 128, shuffle-enabled at the participant level, and the model with the best validation-loss was selected.

### Phase 2: Algorithm Deployment

The use case for deploying the model developed in Phase 1 relates to identifying fall injuries leading to medical attention in Medicare data. Historically, fall injuries leading to medical attention have been identified via participant self-report (usually involving phone calls to participants or postal questionnaires) and/or through full-text review of medical records by trained clinicians.^13^ Gathering data through these processes can be labor-intensive and thus costly, and self-reported data may be incomplete due to participant loss to follow-up^14^ or participants not recalling their events.^15^ If fall injuries can be accurately identified through routinely collected data in administrative databases like Medicare, observational and interventional studies could be conducted at lower cost.

In previous work, we showed that a rule-based algorithm using ICD-10-CM and CPT codes from Medicare data could detect fall injuries leading to medical attention with reasonable accuracy, using self-reported data as the external reference standard (area under the receiver operating characteristic curve [AUC], 0.79, 95% confidence interval, 0.78-0.81).^4^ We hypothesized that the transformer model might perform better than a rule-based algorithm for several reasons. First, the transformer model could potentially be more tolerant to variations in coding practice within administrative data, where non-standard but conceptually similar codes might be used to describe a fall injury (e.g., ICD-10-CM code R22.0, “Localized swelling, mass and lump, head” in lieu of S00.03, “Contusion of scalp”). Second, the transformer model might have improved accuracy by accounting for implicit associations with other codes that are often used in the context of a fall injury but are not diagnostic (and thus not included in the rule-based algorithm) (e.g., ICD-10-CM code R26, “Abnormalities of gait and mobility”). Third, the ability to semantically map codes using the embedding of the text descriptions might improve accuracy. Fourth, the transformer model’s ability to provide a continuous prediction between zero and one could allow for more flexibility to optimize algorithm sensitivity versus specificity (depending on the needs of the use case) by adjusting the probability threshold for identifying a fall injury in the data.

### Phase 2 Evaluation

The phase 2 evaluation included a) checking for general convergence of transformer model ensemble results with the rule-based algorithm and b) comparing the performance of the transformer model ensemble against the same external reference standard that was used for the rule-based algorithm in our prior work (self-reported fall injuries leading to medical attention).^4^ The dataset for the phase 2 evaluation was the full dataset from the STRIDE study, including linked Medicare claims and encounters (n=4895 participants with at least one claim or encounter). To check for general convergence of results with the rule-based algorithm, we looked at the transformer model ensemble’s predictions on a) days when the rule-based algorithm identified the starting day of a new fall injury episode and b) all other days. Since the transformer model evaluates outcomes at the level of the individual claim or encounter, and there can be multiple claims or encounters per day, we took the maximum daily value across all of the mean claim/encounter-level predictions from the 10-model ensemble to be the relevant value for the day, reasoning that not all claims or encounters on a given day might be for a fall injury; thus, the maximum value represents the most valid “signal” of an event on that day. For diagnostic purposes, these predictions were plotted as residuals against the rule-based algorithm’s binary results (“1” for the starting day of a fall injury episode, “0” otherwise).

To compare the transformer model ensemble against the external reference standard, we calculated AUCs. Since dates from the reference standard were self-reported by study participants and thus subject to error, we segmented follow-up time for each participant into “date windows” to compare results of the transformer model ensemble against the reference standard. Date windows could be as narrow as one day (essentially comparing the maximum of the predicted mean value from the model ensemble for the day against whether a fall-injury leading to medical attention was reported on that exact day) or as wide as the entire follow-up period available for each participant (in which case the maximum predicted mean value from the model ensemble for the entire follow-up period was used), with intermediate-sized date windows of 7, 15, 30, 60, 90, 120, 240, 360, and 720 days.

As an exploratory analysis, we also created receiver operating characteristic curves to allow a direct comparison with the rule-based algorithm, using the same methods as in our prior work.^4^ To do this, we had to impose two additional rules on the transformer model predictions. First, since the ultimate goal is to identify unique fall injuries rather than individual claims/encounters for fall injuries, and because care (and thus claims/encounters) for a fall injury may take place over a period of time (e.g., days, weeks, or months [ranging from the initial injury date]), we created episodes of care starting from the earliest matching event date. We tested three different episode lengths (45 days, 90 days, and 180 days). No new fall injury events are identified until the episode of care is completed. Second, we needed to choose a probability threshold above which a claim or encounter on a particular date would be considered a fall injury event. We initially explored thresholds of 0.5, 0.7, and 0.9, and subsequently added 0.95, and then 0.925. We found that an episode length of 90 days and a threshold of 0.925 produced event rates most closely matching the reference standard. We then created receiver operating characteristic curves for the data as a whole, as well as stratified by data source (traditional Medicare or Medicare Advantage) and by the ten healthcare systems participating in the STRIDE study.

## RESULTS

### Descriptive results: Phase 1

We first describe findings from the annotated dataset used to train and validate the transformer model. Across the ten folds of data, on average about 3 ICD-10-CM codes and 1.5 CPT codes were embedded per claim/encounter (Table 1), with a modal value of 1 ICD-10-CM and 1 CPT code (Figure 3), but with significant variation, reflecting the skewed nature of the data.

**Figure 3.**
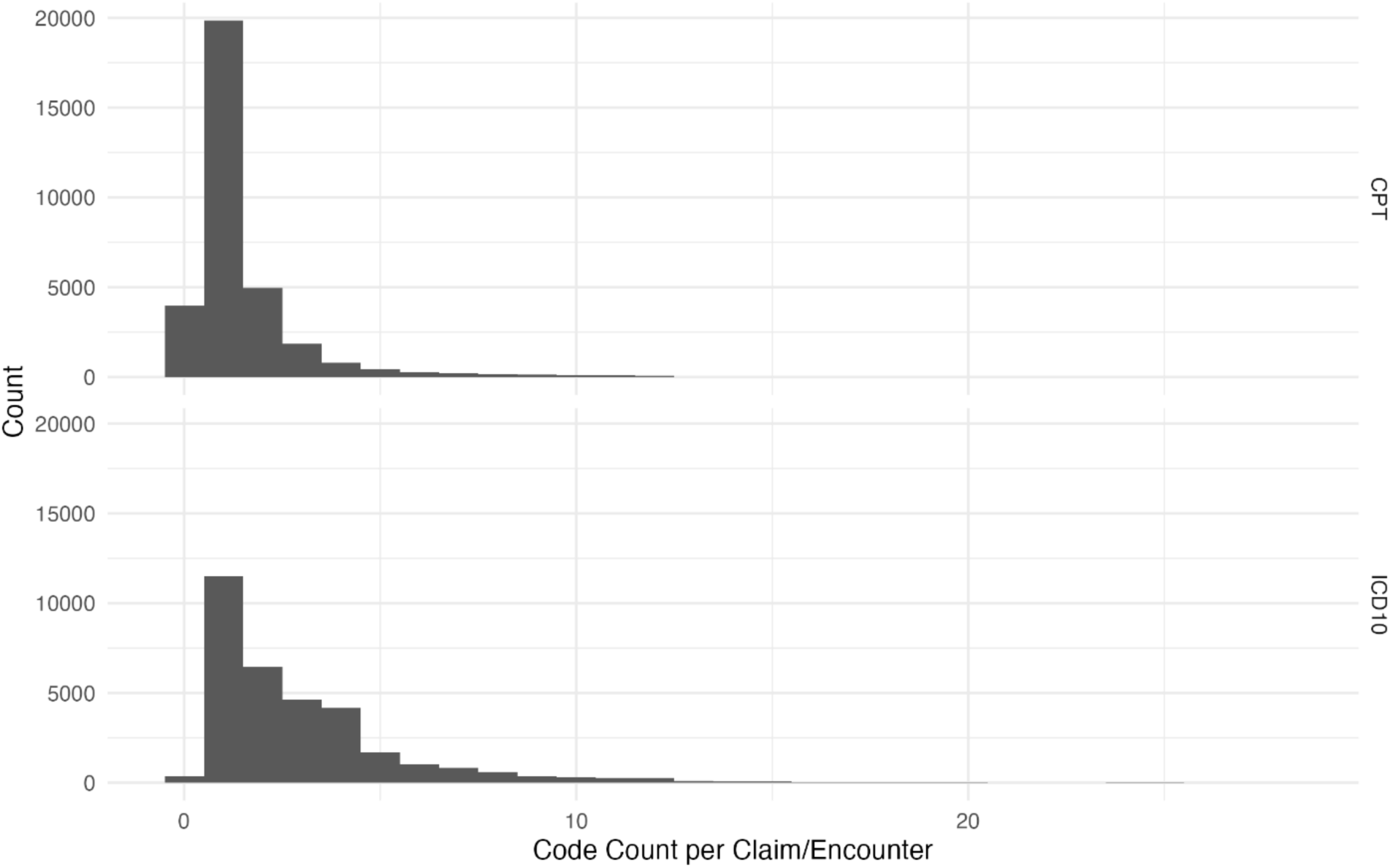
Histogram showing the number of embedded ICD-10 and CPT codes per claim/encounter (row of data)

**Table 1.** Descriptive statistics of available and embedded codes per claim, by fold.

| **Fold** | **ICD-10 total**<br>**mean +/- SD** | **ICD-10 embedded**<br>**mean +/- SD** | **HCPCS (including CPT) total**<br>**mean +/- SD** | **CPT embedded**<br>**mean +/- SD** |
| --- | --- | --- | --- | --- |
| **1** | 3.27 ± 3.09 | 3.16 ± 3.04 | 1.96 ± 1.82 | 1.53 ± 1.46 |
| **2** | 3.09 ± 2.84 | 3.00 ± 2.79 | 1.94 ± 1.95 | 1.49 ± 1.59 |
| **3** | 3.28 ± 2.96 | 3.16 ± 2.89 | 1.96 ± 1.92 | 1.53 ± 1.56 |
| **4** | 3.06 ± 2.78 | 2.95 ± 2.73 | 1.96 ± 1.94 | 1.56 ± 1.67 |
| **5** | 3.02 ± 2.87 | 2.93 ± 2.81 | 1.96 ± 2.01 | 1.53 ± 1.61 |
| **6** | 3.27 ± 3.07 | 3.13 ± 2.99 | 1.92 ± 1.91 | 1.46 ± 1.60 |
| **7** | 3.08 ± 2.85 | 3.00 ± 2.78 | 1.91 ± 1.81 | 1.50 ± 1.56 |
| **8** | 3.42 ± 3.34 | 3.29 ± 3.25 | 1.95 ± 2.05 | 1.43 ± 1.57 |
| **9** | 3.01 ± 2.72 | 2.91 ± 2.67 | 1.90 ± 1.86 | 1.48 ± 1.60 |
| **10** | 3.23 ± 3.02 | 3.11 ± 2.93 | 1.92 ± 1.95 | 1.50 ± 1.58 |

This table shows the total number of codes available per annotated claim, along with the actual number of codes embedded. In the case of ICD-10 codes, some may not have been embedded due to differences between the available codes used in the calendar years of data (2015-2019) versus the year of codes that were embedded (2022). In the case of HCPCS codes, only the subset of HCPCS codes that were part of CPT (2018) were embedded, since the algorithm focused on using CPT codes. Abbreviations: ICD-10, International Classification of Diseases, 10<sup>th</sup> Revision; HCPCS, Healthcare Common Procedure Coding System; CPT, Current Procedural Terminology.

### Phase 1: Model Performance

Indices of model performance are shown in Supplement Figure 1 (absolute residual of model prediction minus annotated value) and Table 2 (model performance in the 10 holdout folds). The model demonstrated an AUC ≥ 0.96 in 9 out of 10 folds and 0.85 in the remaining fold.

**Table 2.** Indices of model performance.

| Fold | Participant N | Training N (rows of data) | Accuracy | Balanced accuracy | Sens. (recall) | Spec. | PPV (precision) | NPV | $\kappa$ | AUC |
| --- | --- | --- | --- | --- | --- | --- | --- | --- | --- | --- |
| 1 | 138 | 3430 | 0.97 | 0.97 | 0.96 | 0.97 | 0.99 | 0.92 | 0.92 | 0.98 |
| 2 | 138 | 3489 | 0.98 | 0.98 | 0.98 | 0.98 | 0.99 | 0.96 | 0.96 | 0.99 |
| 3 | 138 | 3604 | 0.76 | 0.77 | 0.76 | 0.78 | 0.88 | 0.60 | 0.50 | 0.85 |
| 4 | 138 | 2951 | 0.97 | 0.97 | 0.98 | 0.95 | 0.98 | 0.95 | 0.93 | 0.98 |
| 5 | 137 | 3215 | 0.96 | 0.97 | 0.95 | 0.98 | 0.99 | 0.91 | 0.92 | 0.98 |
| 6 | 137 | 3218 | 0.95 | 0.96 | 0.95 | 0.97 | 0.99 | 0.89 | 0.89 | 0.98 |
| 7 | 137 | 3290 | 0.95 | 0.96 | 0.94 | 0.98 | 0.99 | 0.87 | 0.89 | 0.98 |
| 8 | 137 | 3257 | 0.95 | 0.96 | 0.95 | 0.97 | 0.98 | 0.90 | 0.90 | 0.96 |
| 9 | 137 | 3398 | 0.96 | 0.96 | 0.95 | 0.97 | 0.98 | 0.90 | 0.90 | 0.98 |
| 10 | 137 | 3065 | 0.95 | 0.96 | 0.94 | 0.97 | 0.98 | 0.91 | 0.90 | 0.97 |

This table shows indices of model performance by fold of data. Each of the 10 folds shown here represents the “holdout” fold that was not used for model training or validation. Abbreviations: Sens., sensitivity; Spec., specificity; PPV, positive predictive value; NPV, negative predictive value; $\kappa$ , Kappa; AUC, area under the receiver operating characteristic curve.

### Phase 2 analyses

Figure 4 shows the comparability of the text-embedded transformer model predictions against binary results for the rule-based algorithm, at the level of a calendar day. These histograms show general concordance between the transformer model ensemble and the rule-based algorithm. Figure 5 shows example data plotted at the calendar-day level for four exemplar cases, showing both concordant and discordant scenarios when comparing the transformer model with the rule-based algorithm.

**Figure 4.**
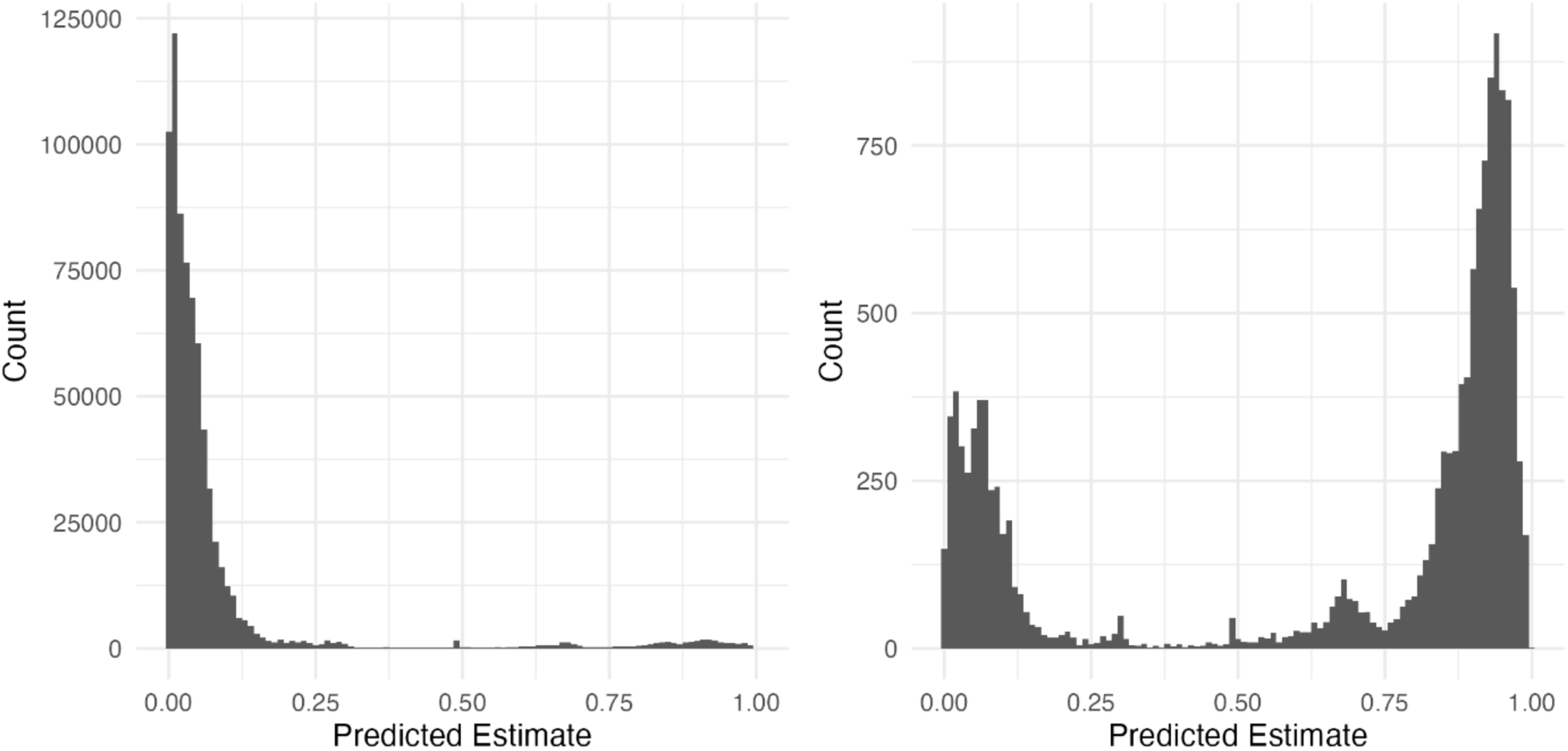
Histograms comparing transformer model predictions against rule-based algorithm results. Figure 4a (left-hand panel) is a histogram of the maximum of the predicted mean probability of a fall injury on a given date from the transformer model when a rule-based algorithm to identify fall injuries showed the date as <u>not</u> an index date for an injury episode. Figure 4b (right-hand panel) is a histogram of the maximum predicted mean probability of a fall injury on a given date from the transformer model when a rule-based algorithm did identify the date as an index date for a full injury episode. Note the difference of scale on y-axes for Figure 4a and Figure 4b.

**Figure 5.**
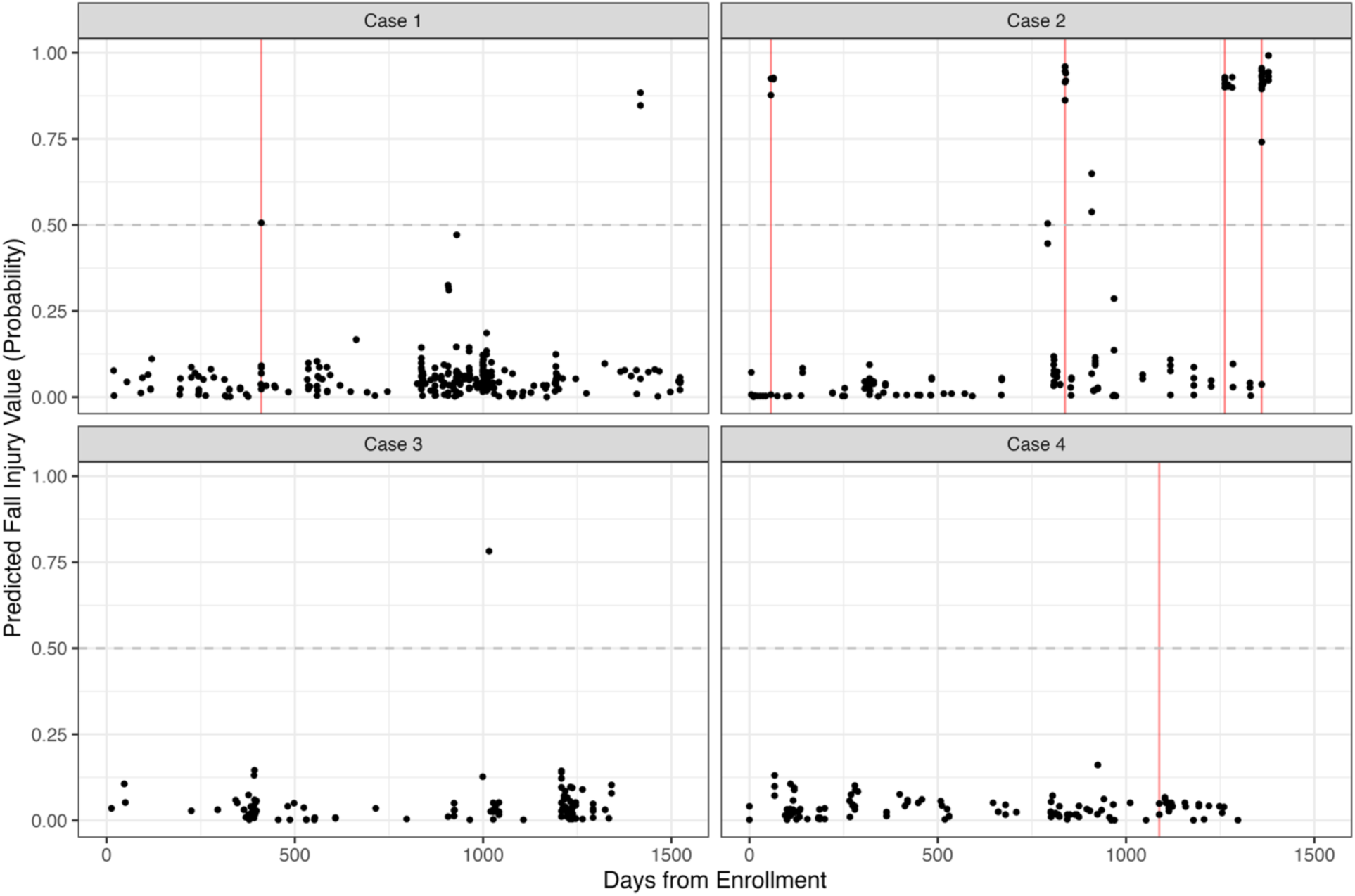
Depictions of four exemplar cases. Figure 5 shows four exemplar cases (participants). The y axis shows the predicted probability of a fall injury based on the transformer matching model, and the x axis represents the elapsed time (in days) from enrollment in the study. Each dot represents a particular claim or encounter for healthcare. The dashed line demarcates a probability of 0.5; dots that fall above the line are ones in which the transformer model predicts that an event is more likely than not. Red lines represent a day on which the rule-based algorithm indicated the start of an episode of care for a fall injury. In Case 1 (upper left panel), the rule-based algorithm identified one event during follow-up, for which the transformer model found one claim/encounter with a probability of about 0.5. The transformer model identified another high-probability event later in follow-up for Case 1. In Case 2 (upper right panel) there was evidence of general concordance between the transformer model and the rule-based algorithm across four events. In Case 3 (lower left panel), the transformer model identified an event with a probability of about 0.75 that was not identified by the rule-based algorithm. In Case 4 (lower right panel), the rule-based algorithm identified an event that was not identified by the transformer model.

Table 3 shows transformer model performance against the external reference standard of self-reported fall injuries leading to medical attention, with inexact matching on dates being allowed using “date windows.” AUC remained at 0.83 or higher for date windows of 30 days or longer, with peak accuracy achieved using a date window of 60 days (AUC 0.86; 95% CI, 0.84-0.87).

**Table 3.** Area under the receiver operating characteristic curve for the transformer model ensemble compared to the reference standard of self-reported fall injuries leading to medical attention.

| window size | AUC (95% CI) |
| --- | --- |
| 1 day | 0.68 (0.67 – 0.69) |
| 7 days | 0.77 (0.76 – 0.78) |
| 15 days | 0.80 (0.79 – 0.82) |
| 30 days | 0.83 (0.82 – 0.85) |
| 60 days | 0.86 (0.84 – 0.87) |
| 90 days | 0.85 (0.84 – 0.86) |
| 120 days | 0.85 (0.84 – 0.87) |
| 240 days | 0.85 (0.84 – 0.87) |
| 360 days | 0.84 (0.83 – 0.86) |
| 720 days | 0.83 (0.81 – 0.85) |
| FULL follow-up period | 0.85 (0.83 – 0.86) |

For each row in this table, we varied the probability threshold for model event detection between zero and one, with self-reported fall injuries leading to medical attention serving as the reference standard. Each row in the table represents a different analysis, varying the stringency of required date match between self-report and the machine learning algorithm from one day (most stringent), to the full period of follow-up for each participant (least stringent). For example, for a date window of 30 days, at any given probability threshold for the model, sensitivity was calculated as the proportion of date windows with one or more self-reported fall injuries leading to medical attention in which the maximum predicted mean probability of the model ensemble during the date window exceeded the given threshold; specificity was calculated as the proportion of date windows with zero self-reported fall injuries leading to medical attention in which the maximum predicted mean probability of an event derived from the model ensemble remained under the given threshold. Varying the given probability from zero to one generated the AUC result for the 30-day date window shown above. Abbreviations: AUC, area under the receiver operating characteristic curve.

Figure 6 shows transformer model performance alongside rule-based algorithm performance after choosing a probability threshold for the transformer model (probability = 0.925) as well as the length of an episode of care after a fall injury (90 days) to allow for side-by-side comparison. The transformer model ensemble performed comparably to the rule-based algorithm, both on the full dataset (AUC=0.79) as well as by data type (fee-for-service Medicare claims versus Medicare Advantage encounters) and by the ten participating healthcare systems in the study.

**Figure 6.**
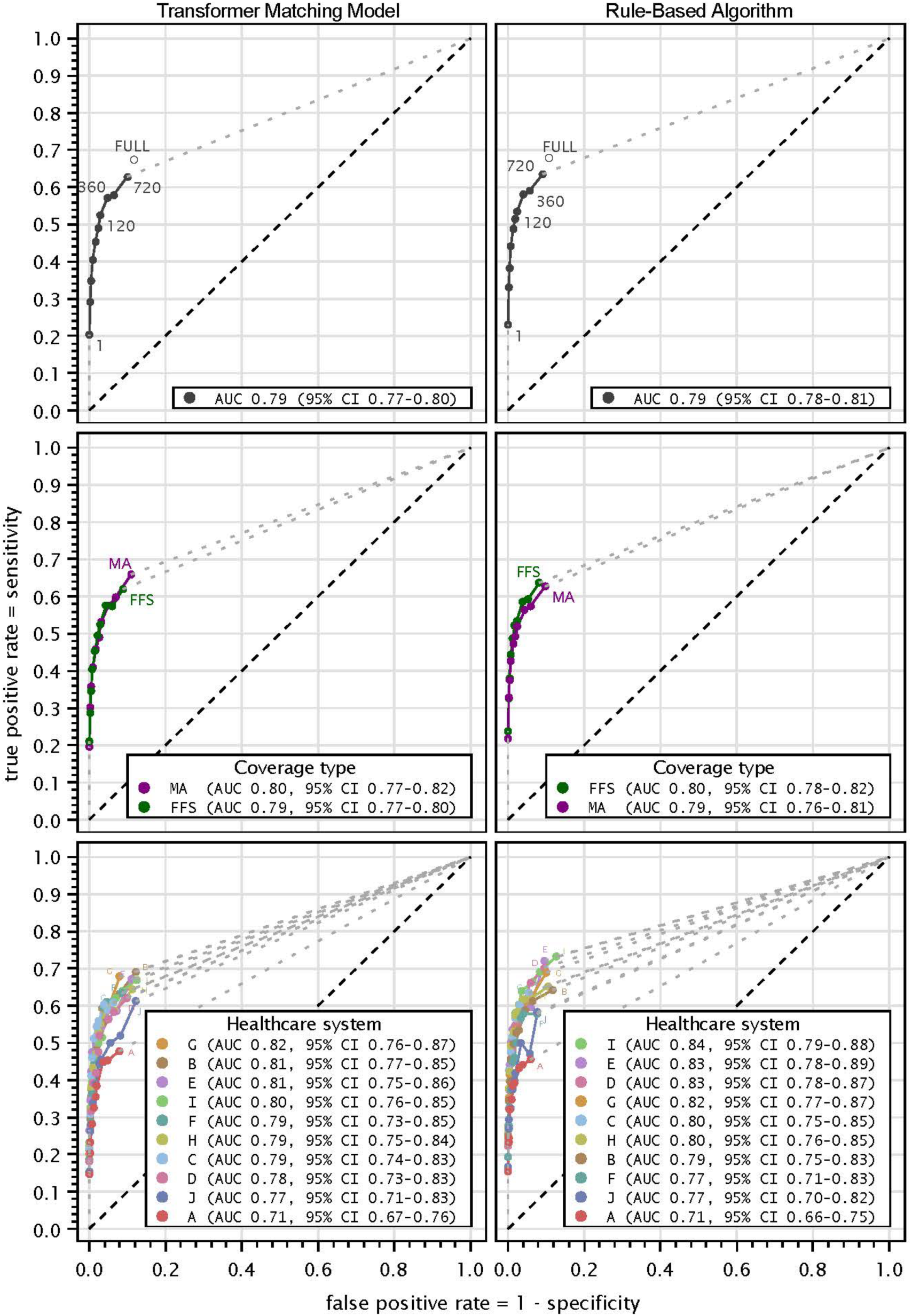
Comparison of transformer model and rule-based algorithm performance. Figure 6 shows area under the receiver operating characteristic curve compared with an external reference standard (self-reported fall injuries leading to medical attention). The left column shows results of the transformer matching model ensemble LLM, while the right column shows results of the rule-based algorithm. Within the figure, there are six panels. The y axis of each panel shows the true positive rate (sensitivity), and the x-axis shows the false positive rate (1-specificity). Inset boxes provide the area under the receiver operating characteristic curve and 95% confidence limits. The gray dashed lines demarcate the full receiver operating characteristic curve. The top row shows the overall results; numbers shown by each point represent the length of the date window in days over which the true positive rate and false positive rate were calculated, ranging from 1 day to 720 days. “FULL” represents a date window representing the full duration of follow-up for each participant; this is not part of the curve. The middle row shows results stratified by Medicare Advantage versus Medicare fee-for-service (among individuals continuously enrolled in Medicare Advantage or fee-for-service Medicare); the bottom row shows results stratified by the ten healthcare systems that participated in the study. AUC = area under the receiver operating characteristic curve; CI = confidence interval; FFS = Medicare fee-for-service; MA = Medicare Advantage.

## DISCUSSION

We found that a supervised text-embedded transformer matching model could achieve performance comparable to a rule-based algorithm for detecting fall injuries in Medicare data. The use of a transformer model in combination with LLM embeddings represents a relatively new approach to handling structured data such as ICD-10-CM and CPT codes. Kirchler and colleagues described GRASP, which similarly used LLM embeddings from structured terminologies (in their case, Observational Medical Outcomes Partnership [OMOP] vocabulary concepts) as inputs into a transformer-based model to predict a variety of diagnoses.^16^ Our dataset is unique in having an external (completely independent) reference standard in the form of self-reported fall injuries leading to medical attention, which allowed us to both annotate the claims and encounter data for training purposes and estimate algorithm performance as compared with a rule-based algorithm.

Contrary to our initial expectations, we found that the text-embedded transformer model did not outperform the rule-based algorithm when held to comparable standards. Based on the experience of annotating the raw data, in which we observed many instances of non-specific coding being used (e.g., T14.90, “Injury, unspecified”), we speculate that limitations in coding quality in the underlying data impose an upper limit on the potential performance of any model. (In addition, in the specific instance of this work, the imperfect nature of the self-reported reference standard, which is subject to the limitations of human recall, may also cause a decrement in observed performance.) In the scenario where variable coding quality is the norm, we see the transformer model as offering an alternative approach that may make it more useful in some circumstances, while the rule-based algorithm may be more useful in others. In clinical trials, the transformer model could be used as part of a hybrid approach in which some events (e.g., probability of at least 0.925) are automatically confirmed, while other events (e.g., probability of at least 0.5 but less than 0.925) are manually reviewed and adjudicated with full text of medical records if needed. In contrast, the rule-based algorithm has high specificity but only moderate sensitivity, with no opportunity to adjudicate “possible” events (since these are not identified by the algorithm). However, the rule-based algorithm can provide detailed information about the type of injury and duration of the episode of care that is not yet possible from the transformer model.

Our work has certain limitations. First, the training, validation, and holdout datasets used in the initial development of the transformer model were a subset of the full data in which the transformer model was ultimately deployed for comparison against the rule-based algorithm. For this reason, performance of the transformer model in the full dataset may be optimistic relative to a completely new sample. Second, the use case of identifying fall injuries in Medicare data represents a very specific type of event whose identification may be favored by the acuity of these events and the detailed nature of ICD-10-CM coding for injuries in particular. Third, our data derived from a clinical trial whose sites’ coding patterns may or may not be representative of the general coding patterns present in Medicare data. For this reason, future work should reverify performance in a nationally representative sample.

Logical extensions of the current work include the ability to develop refined event predictions by adding new sources of data. For example, adding participant-level demographics (e.g., age, sex) could improve fit. In addition, a combined model that uses both structured data from claims and encounters along with full text of clinical notes documented in electronic health records could improve model performance. A model drawing on both structured data and unstructured clinical notes could be tested against traditional adjudication approaches, such as has been explored in the cardiovascular literature.^17,18^

## CONCLUSION

We demonstrated the feasibility of using a text-embedded transformer model to predict fall injuries in Medicare data and the ability to achieve comparable results as compared with a rule-based algorithm.

## ACKNOWLEDGEMENTS

The organizations funding this study had no role in the design or conduct of the study; in the collection, management, analysis, or interpretation of the data; or in the preparation, review, or approval of the manuscript. Artificial intelligence (Claude Code [Anthropic]) was used to assist in generation of Figures 1 and 2. The content of this publication is solely the responsibility of the authors and does not necessarily represent the official views of the National Institutes of Health, the Department of Veterans Affairs, or the United States government.

## AUTHOR CONTRIBUTIONS

MK and DAG conceptualized and designed this study. DAG, DE, and NKL were responsible for acquisition of data. MK, EJG, and DAG analyzed the data; all authors interpreted the data. MK and DAG drafted the manuscript, and all authors provided critical revisions to the manuscript for important intellectual content. All authors approved the submitted manuscript.

## DATA AVAILABILITY

Due to the terms of the data use agreement with the US Centers for Medicare and Medicaid Services, Medicare data cannot be shared with third parties. A deidentified version of the original STRIDE study data is available through a request process at the Aging Research Biobank (https://agingresearchbiobank.nia.nih.gov/).

## Supplement

**Supplement Figure 1.**
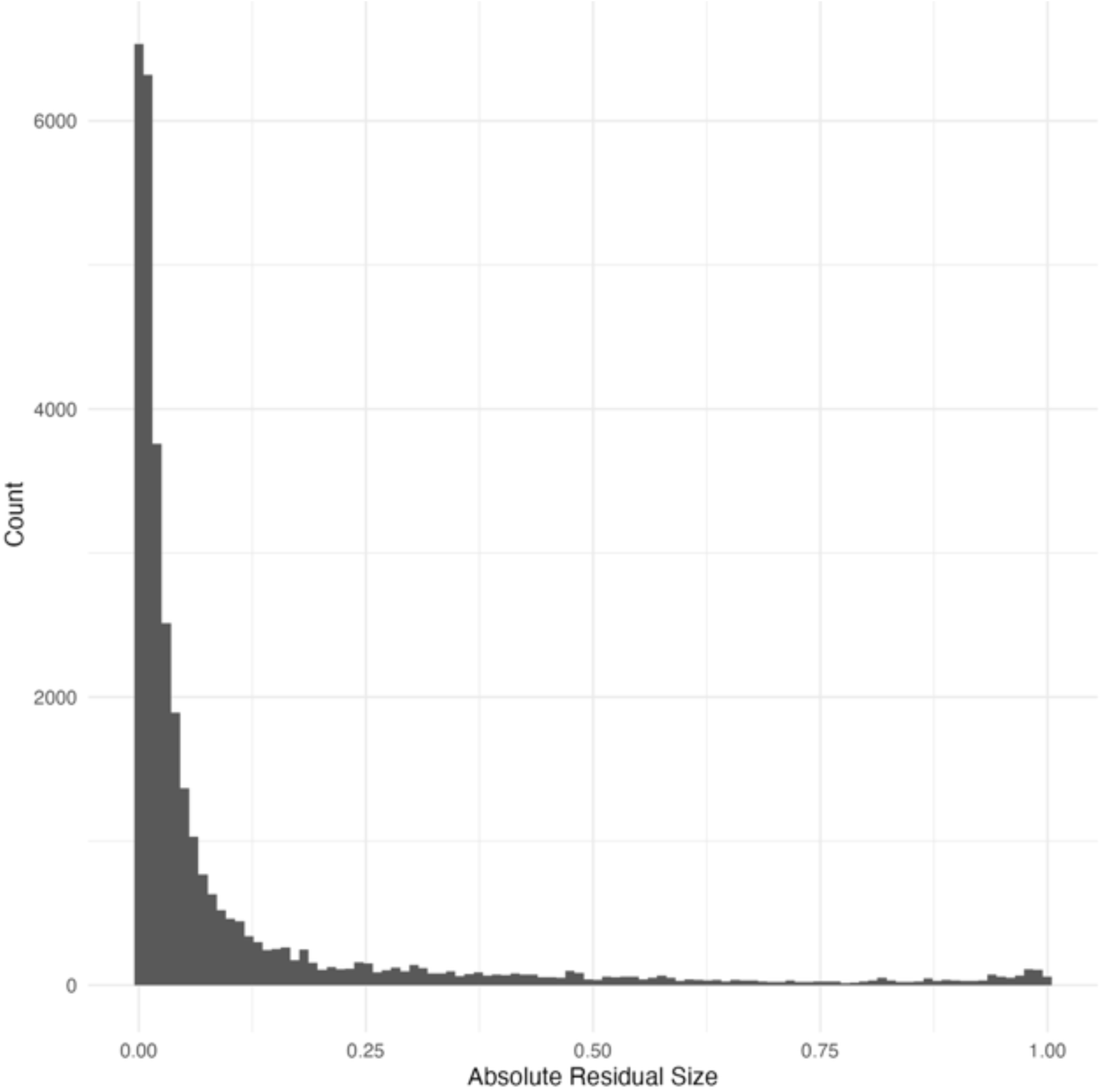
Histogram of absolute residual size (absolute value of model prediction minus annotated result) for each claim/encounter (row of data, N=32,917) Supplement Figure 1 depicts the absolute residual size, which is the absolute value of the difference between the model’s mean predicted probability of being a claim for a fall injury (mean value across all ten models [one for each holdout fold]), and the annotated result (0 or 1).

